# Hepatitis C subtyping assay failure in UK patients born in Sub-Saharan Africa: implications for global treatment and elimination

**DOI:** 10.1101/2021.12.14.21267714

**Authors:** Kazeem Adeboyejo, Barnabas J. King, Theocharis Tsoleridis, Alexander W. Tarr, John McLauchlan, William L. Irving, Jonathan K. Ball, C. Patrick McClure

## Abstract

The newly developed direct-acting antivirals (DAAs) have revolutionised the treatment of chronic hepatitis C virus (HCV), where cohort studies have shown that cure rates as high as 98% can be achieved. Whilst genome sequencing has demonstrated that some subtypes of HCV naturally harbour drug resistance associated substitutions (RAS), these have not been considered important as previous molecular epidemiological studies have suggested that such difficult-to-treat subtypes are rare. Therefore, to optimise and streamline molecular detection and sequence-based typing of diverse RAS-containing subtypes, a novel panel of single round PCR assays was applied to HCV derived from 146 individuals, whose likely source of infection was from regions of sub-Saharan Africa (SSA). Partial NS5A and NS5B sequences were obtained from 135 HCV-positive patients born in 19 different countries from SSA but attending clinics in the UK.

Virus subtype assignments were determined by pairwise-distance analysis and compared to both diagnostic laboratory assignments and free-to-use online typing tools. We determined that routine clinical diagnostic methods incorrectly subtyped 59.0% of samples, with a further 6.8% incorrectly genotyped. Of five commonly used online tools, Geno2Pheno performed most effectively in determining a subtype in agreement with pairwise distance analysis. Considering the estimated number of HCV infections to have occurred in across Africa, this study provides a simple low-cost pathway to guide regional therapeutic choice and assist global eradication programmes.

## Introduction

The World Health Organisation’s (WHO) global plans to eliminate hepatitis C virus (HCV) as a public health threat, detailed in 2016 [1], placed a significant dependency upon the use of the new generation of interferon-free direct-acting antiviral (DAA) drug regimens. In the five years since the release of that report there has been a steady increase in the reporting of natural resistance-associated substitutions (RAS) in patients who acquired the virus in previously under-surveyed areas, such as countries in sub-Saharan Africa (SSA) [2-4] and South America [5, 6].

According to United Nations data, as of 2019, an estimated 1.1 billion people live in sub-Saharan Africa [7]. Whilst estimates of HCV seroprevalence in the region vary greatly [8-11], a recent systematic review of a large number of studies estimated the number of people infected is approximately 37 million – around 20% of the global HCV burden [12]. A number of molecular epidemiological studies have shown that the region contains a highly diverse population of HCV, with the reported dominating genotypes varying significantly according to geographical location and study [13-22].

Assignment of HCV strains into genotypes and subtypes is done according to criteria set by the International Committee on Taxonomy of Viruses [23]. HCV genotypes differ by >30% and subtypes by >15% [24] across the complete protein-coding region. While defining new subtypes requires >95% of the coding region of an isolate to be sequenced; for clinical and epidemiological purposes a smaller region of the genome, typically of the NS5B gene, is used [25].

With the recent advent of DAAs, accurate subtype identification has become more important, as different genotypes and subtypes respond with varying degrees of success to DAA-based regimens [26]. For example, recent UK studies of DAA treatment failure (including an NS5A-targeting antiviral) in individuals originating from SSA demonstrated that the patients were infected with subtypes Gt1l and Gt4r, which encode naturally occurring RAS to NS5A-targeting drugs, such as ledipasvir (LDV) or daclatasvir (DCV) [22, 27]. Furthermore, a review of multiple studies observed that 8-16% of patients with Gt1 HCV infections had naturally occurring NS5A RAS that could impact ledipasvir-based treatments [28]. The apparent low prevalence of subtypes harbouring RAS in developed countries, where DAA therapy is more widely used, has led to several authors referring to these resistant-subtypes as ‘rare’ [29]. However, emerging molecular epidemiological evidence suggests that in some parts of SSA, for example Ethiopia and Uganda, DAA-RAS-associated subtypes are relatively frequent [2-4]. If these trends prove broadly representative of SSA in general, then this will have a major impact on first line therapies and HCV treatment and eradication not only in SSA, but also countries with significant ex-patriot SSA populations [22].

The treatments currently approved for use in the UK are: sofosbuvir; ombitasvir-paritaprevir-ritonavir-(dasabuvir); ledipasvir-sofosbuvir; elbasvir-grazoprevir; sofosbuvir-velpatasvir; glecaprevir-pibrentasvir and sofosbuvir-velpatasvir-voxilaprevir [30]. A recent study of patients who failed DAA therapy, the majority of whom had received sofosbuvir-based therapy, revealed subtype-specific RAS [31] underscoring the need for accurate subtyping. There is very limited evidence regarding the efficacy of these regimens against more recently described subtypes [32], many of which have naturally occurring pre-treatment DAA-RAS.

Due to the increased subtype diversity, patients who are likely to have acquired HCV in SSA, attending hepatology clinics in the UK represent a particular challenge in treatment selection because treatment is not exclusively pan-genotypic and is thus frequently based upon virus genotype/subtype, necessitating accurate diagnostic typing tools. In this study, we developed a novel panel of robust PCR primers to efficiently obtain partial NS5A and NS5B sequence data but sufficient for typing and resistance profiling from 135 HCV-positive patients suspected of acquiring infection in SSA but attending clinics in the UK. Virus subtype assignments were determined by phylogenetic analysis and compared to results using freely accessible online tools. These data highlight the difficulties in correctly assigning HCV subtype, which in turn can have serious implications for appropriate first-line therapy choice and planned global elimination.

## Materials and methods

Samples were obtained from HCV Research UK - a cohort of >11,000 patients with chronic HCV infection attending health clinics in the UK and enrolled between 2012-16. Serum was obtained from 146 patients who were HCV RNA-positive, born in SSA and who had not been identified as people who inject drugs (PWID). Having no reported risk-factors for HCV transmission since arrival, this group were suspected to have acquired infection prior to moving to the UK. The Republic and the Democratic Republic of the Congo were not distinguished in the HCV Research UK cohort records. This sample cohort has been extensively described elsewhere [22]

RNA was extracted from 140 µL of serum from each sample using a MinElute virus spin kit (QIAGEN) according to the manufacturer’s instructions. Extracted nucleic acid was eluted into 50 µl elution buffer and stored at −70°C. 20 µl of nucleic acid extract was added as template for complementary DNA (cDNA) synthesis using lyophilized RNA to cDNA EcoDry^™^ Premix (random hexamers) (Takara-Clontech), according to manufacturer’s instructions, and stored at −20°C. Degenerate primers were used to amplify partial NS5A (genotype-specific primers) and NS5B (pan-genotype primers) regions (**Table 1**). PCR was performed using HotStarTaq DNA polymerase (QIAGEN) in 20 µl reactions containing 1µl cDNA template and 0.5 µM (NS5B, [25]) or 1.0 µM (NS5A) of each primer. PCR reactions were initially heated at 95°C for 15 mins followed by 55 cycles of: 95°C for 20 secs, 56°C for 20 secs, 72°C for 30 secs (NS5B) or 1 min (NS5A) followed by a final extension elongation step of 72°C for 1 min. Products were visualised by agarose gel electrophoresis, and putative positives submitted for Sanger sequencing (Source BioScience, Nottingham, UK) by ten-fold dilution in water. Sanger sequences were manually checked for quality and accuracy using FINCHTV base-calling software and terminal primer sequences removed (GenBank accession numbers, NS5B: MT151021 - MT151155; NS5A: MT151156 - MT151286). Resulting nucleotide data was aligned with ICTV HCV reference sequences and processed with Geneious Prime 2019.0.4 software. A total of 352 sequences were aligned based on their partial 316nt *NS5b* gene using MAFFT. A Maximum-likelihood phylogenetic tree was generated using IQ-TREE2, utilising a GTR+F+R10 model of nucleotide substitution as suggested by the software’s model finder, with 1000 SH-like approximate likelihood ratio test (SH-aLRT). Phylogenetic trees were annotated using FigTree v.1.4.4.

**Table 1.**
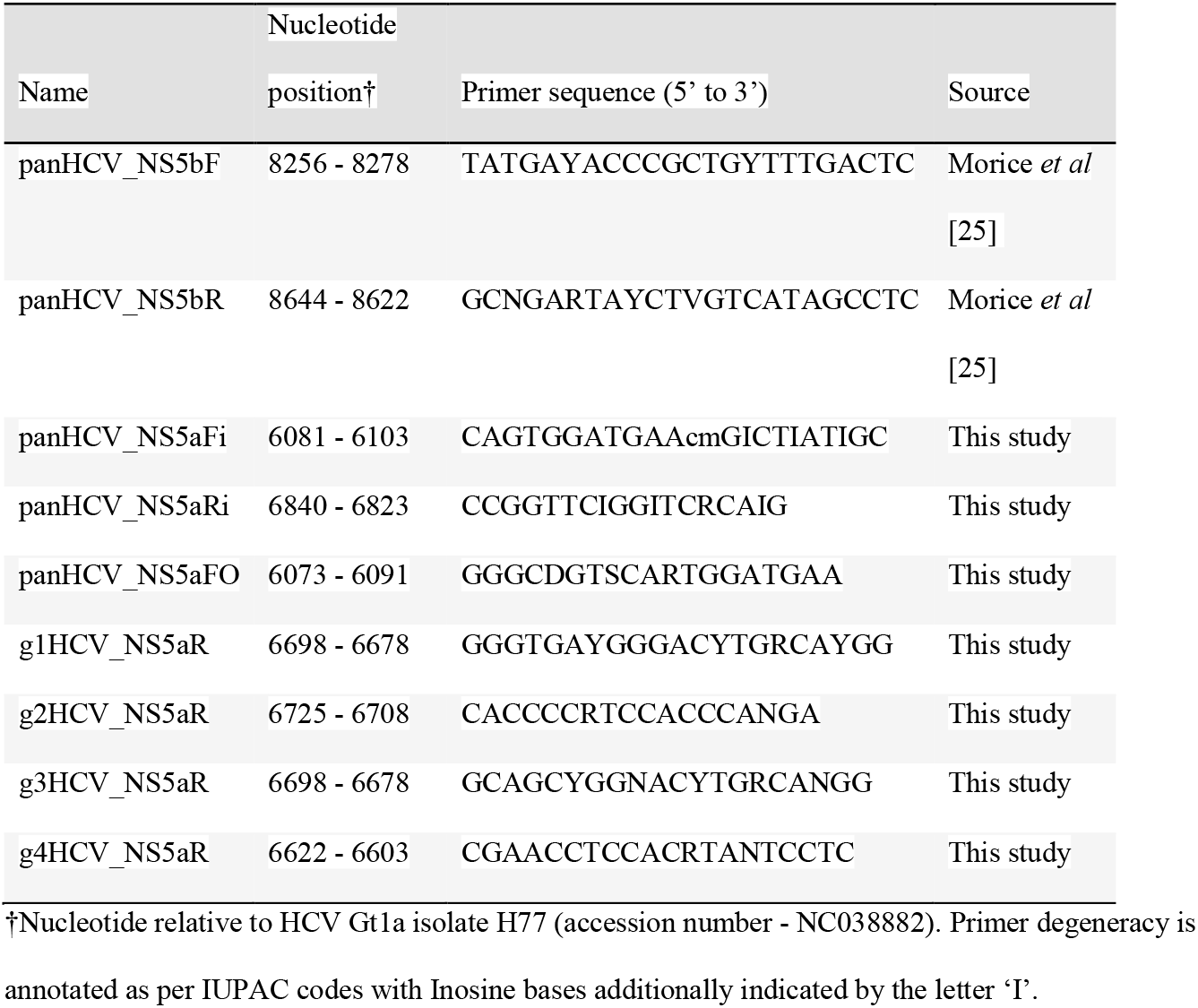
Primers used in this study

HCV subtyping was performed by comparative analysis with isolates in a reference dataset (International Committee on Taxonomy of Viruses [ICTV] Confirmed HCV genotypes/subtypes [May 2019] (https://talk.ictvonline.org/ictv_wikis/flaviviridae/w/sg_flavi/57/hcv-reference-sequence-alignments) using an uncorrected pairwise distance matrix performed on nucleotide sequence alignments trimmed to the respective partial NS5A or NS5B amplicon, using MEGA X.

NS5B sequences were uploaded to five online HCV subtyping tools: Los Alamos National Laboratory (LANL) HCV database, https://hcv.lanl.gov/content/index or Virus Pathogen Database and Analysis Resource (ViPR), https://www.viprbrc.org/brc/home.spg?decorator=vipr (both performed May 2019); and also Genome Detective, https://www.genomedetective.com/app/typingtool/hcv/; Max Plank Institut Informatik Geno2Pheno [HCV], https://www.hcv.geno2pheno.org/ [33] or HCV-GLUE, http://hcv-glue.cvr.gla.ac.uk/#/home [34] (all three performed November 2021). RAS were identified using Geno2Pheno [HCV].

## Results

### Sample summary

Initially, a partial NS5B fragment was successfully amplified from 135 samples of HCV-positive patients of SSA birth with no history of intravenous drug use (**Supplementary Table 1**). Despite repeated attempts, no HCV sequence could be amplified from 11 other samples (**Supplementary Table 2**). There was no genotype-associated amplification failure (based upon clinical laboratory genotype assignments) and viral load data was only available for two of these samples. These samples were excluded from all further analysis and description.

The majority of the 135 NS5B-PCR-positive samples were from men (56.3%) and, at the time of sample receipt in November 2017, from individuals aged between 50 and 70 years of age (54.8%) (**Figure 1A**). In the majority of cases (75.6%), no risk factors for infection could be identified. The commonest identifiable risk factor prior to emigrating to the UK was having received blood or blood products (17.0%) (**Figure 1B**). Other suspected routes of transmission recorded were having an HCV-positive sexual partner or medical procedures. The samples were from patients born in nineteen SSA countries, with countries from the Eastern Africa region most represented (**Figure 1C**). The majority of patients were from three countries: Somalia (n=29), Nigeria (n=26) and Congo (n=19). Twelve countries were represented by <5 patients each.

**Figure 1:**
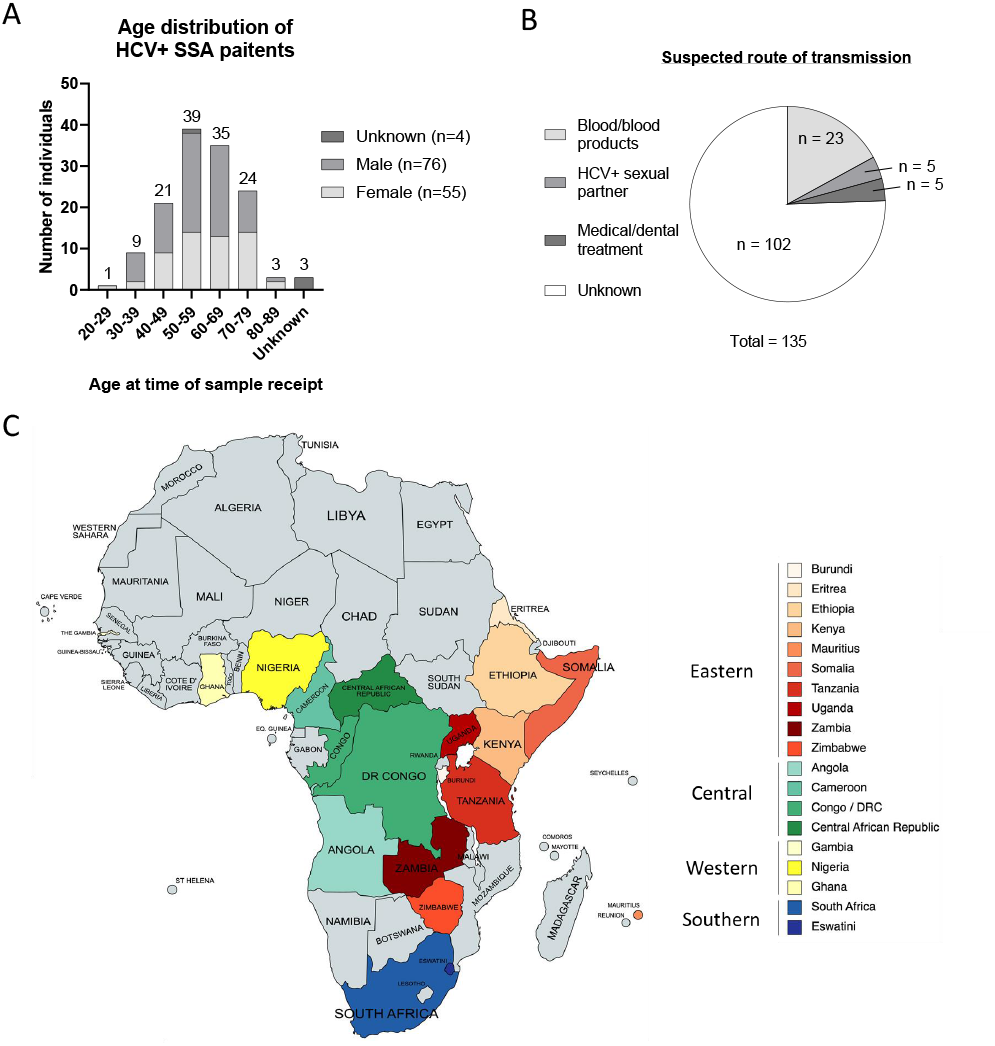
Patient data summary. **A** Age of patients at time of serum-sample receipt from HCV-UK. Date of birth data was no available for 3 patients. Frequency values (n) are shown above columns. **B** The primary suspected route of HCV acquisition for patients in the cohort were detailed. It was not possible to distinguish between patients for whom no data was provided and those where no specific transmission route was suspected. **C** Map of Africa highlighting the country of birth of patients in this study. The number of patients born in each country is shown in brackets. Countries are grouped into African Regions based upon the United Nations Statistics Division (UNSD) classification. It was not possible to distinguish between patients from Democratic Republic of the Congo and Republic of the Congo. These samples were grouped together as ‘Congo’.

### Sample subtyping using NS5B

NS5B sequence data obtained for all the SSA samples were trimmed to 316 bases in length (nucleotides 8305-8620 of H77 reference, accession: AF009606) and assembled into an alignment with a complete 2019 reference dataset produced by the International Committee on Taxonomy of Viruses. A pairwise distance matrix was generated (**Supplementary Table 3**) to describe uncorrected *p*-distances for subtype assignment based upon distance from the closest reference isolate (**Supplementary Table 4**). The subtypes identified by country of patient birth are summarised in **Table 2**.

**Table 2.**
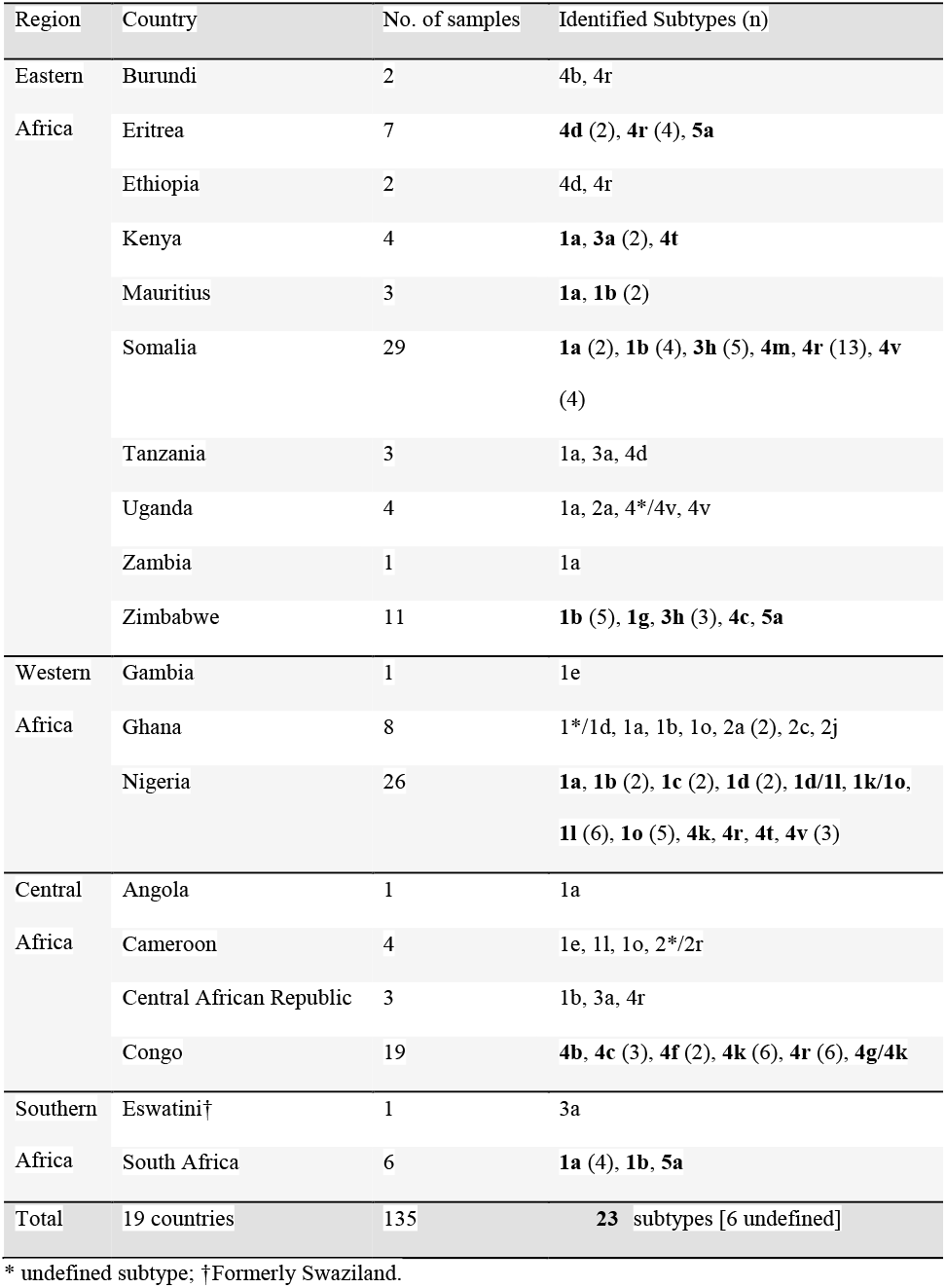
HCV subtypes identified in 135 patients born in sub-Saharan Africa (SSA)

The most common subtype in the cohort was Gt4r (n=27) followed by subtypes Gt1a and Gt1b (14 and 16 samples, respectively) (**Supplementary Figure 1**). Samples were classified into 23 different subtypes with six samples being equally similar to references from two different, or unclassified, subtypes. For the majority of SSA samples the *p-*distance to the closest reference was <0.1 (**Supplementary Table 4**). However, 27 samples had a *p-*distance of ≥0.1 to the nearest reference clone, indicating the need for additional sequencing of full-length clones to allow more accurate sub-typing (**Table 3**). Interestingly, these 27 samples were obtained from individuals from just six areas: Cameroon, Ghana, Nigeria, Uganda, Zimbabwe and the Congo. This may reflect a combination of high diversity and limited detailed characterisation of samples from these countries.

**Table 3.** Samples without a close reference clone (*p*-distance ≥0.1)

| Sample | Subtype using | $p$-distance to closest | Closest reference isolate | | |
| --- | --- | --- | --- | --- | --- |
| ID | partial NS5B | reference† | Reference ID #1 | Reference ID #2 | Reference ID #3 |
| Cam_01 | 2*/2r | 0.14 | >2_JF735119 | >2r_JF735115 |  |
| Con_02 | 4g/4k | 0.13 | >4g_JX227971 | >4k_FJ462438 |  |
| Con_04 | 4b | 0.1 | >4b_FJ462435 |  |  |
| Con_05 | 4f | 0.1 | >4f_EU392175 |  |  |
| Con_09 | 4k | 0.15 | >4k_EU392173 | >4k_FJ462438 |  |
| Gha_01 | 1*/1d | 0.13 | >1_AJ851228 | >1d_KJ439768 |  |
| Gha_02 | 2a | 0.12 | >2a_HQ639944 |  |  |
| Gha_03 | 1o | 0.16 | >1o_KJ439779 |  |  |
| Gha_04 | 1a | 0.14 | >1a_AF009606 |  |  |
| Gha_06 | 2c | 0.12 | >2c_D50409 |  |  |
| Gha_07 | 2a | 0.12 | >2a_HQ639944 |  |  |
| Gha_08 | 2j | 0.12 | >2j_HM777359 |  |  |
| Nig_01 | 1k/1o | 0.14 | >1k_KJ439774 | >1o_KJ439779 | >1o_MH885469 |
| Nig_02 | 1d | 0.12 | >1d_KJ439768 |  |  |
| Nig_09 | 1d/1l | 0.15 | >1d_KJ439768 | >1l_KC248197 |  |
| Nig_10 | 1d | 0.15 | >1d_KJ439768 |  |  |
| Nig_14 | 1o | 0.12 | >1o_KJ439779 |  |  |
| Nig_15 | 1c | 0.11 | >1c_AY051292 | >1c_D14853 |  |
| Nig_18 | 1o | 0.13 | >1o_MH885469 |  |  |
| Nig_20 | 1o | 0.13 | >1o_KJ439779 |  |  |
| Nig_21 | 1c | 0.16 | >1c_AY051292 | >1c_D14853 |  |
| Nig_23 | 1o | 0.15 | >1o_KJ439779 |  |  |
| Nig_25 | 1o | 0.12 | >1o_KJ439779 |  |  |
| Uga_03 | 4*/4v | 0.1 | >4_JF735129 | >4v_HQ537008 | >4v_HQ537009 |
| Zim_03 | 3h | 0.14 | >3h_JF735121 |  |  |
| Zim_07 | 3h | 0.15 | >3h_JF735121 | >3h_JF735126 |  |
| Zim_10 | 3h | 0.16 | >3h_JF735121 | >3h_JF735126 |  |

†Uncorrected *p*-distance, \*undefined subtype

Some patterns of note were observed in the subtypes identified: i) all of the 19 samples from the Congo were Gt4, ii) Gt3h from Zimbabwe was substantially different from the Gt3h reference sequence and the others identified in the cohort, iii) six of the seven Gt1o samples were ≥0.12 divergent from the nearest reference clone, iv) seven of the eight samples from Ghana and 11 of the 26 samples from Nigeria were ≥0.1 from the closest reference isolate.

### Phylogenetic analysis

Investigation of the phylogenetic relationship of the samples by genotype was undertaken alongside the ICTV reference isolates (Supplementary Figures 2A – F, respectively Gt1 to Gt5 individually and all together). Maximum likelihood analysis of the partial NS5B coding nucleotide sequences showed good concordance with the geno- and sub-typing assignments using p-distances. There was good bootstrap support at the genotype level, but less so at the subtype level. Whilst most sequences clustered within known subtypes there were a number that grouped separately, potentially representing unassigned subtypes (Supplementary Figures 2A-F).

### Analysis of diagnostic laboratory genotyping and subtyping accuracy

The HCV Research UK database recorded the assay and primary care centre that conducted the clinical laboratory genotyping and viral load determination; however subtyping information was incomplete. The samples were genotyped using one of seven different methods, including three commercial assay systems (**Supplementary Table 5**). The subtyping assignments made using the *p-* distance matrix were compared to those of the original clinical laboratory subtyping and the outputs of online subtyping resources (**Supplementary Table 5**). Of the 117 diagnostic lab results available, for fifty eight of the samples (49.6%) no subtype was assigned in the HCV Research UK clinical data. Comparing the *p-*distance subtype assignments: 6.8% (n=8) of the samples were mis-genotyped by the clinical lab assays, and 65.8% of samples (total: n=77) were either not assigned (n=58) or were mis-assigned a subtype (n=19) (**Table 4 and Supplementary Table 5**). No commercial or in-house assay showed complete concordance with the p-distance-assigned subtypes. For example, of the three commercial assays used (Abbott, Roche and Siemens) the Siemens assay had the highest concordance with the p-distance assigned subtypes (51.5%). However, the Siemens assay was used on more Gt1a and Gt1b samples (11), compared to the Roche (4) and Abbott assays (5). This is relevant as these genotypes are highly prevalent in the USA and the UK where these assays were developed. There was no consistent assay-breakdown with relation to a given genotype or subtype (**Supplementary Table 5**).

**Table 4.**
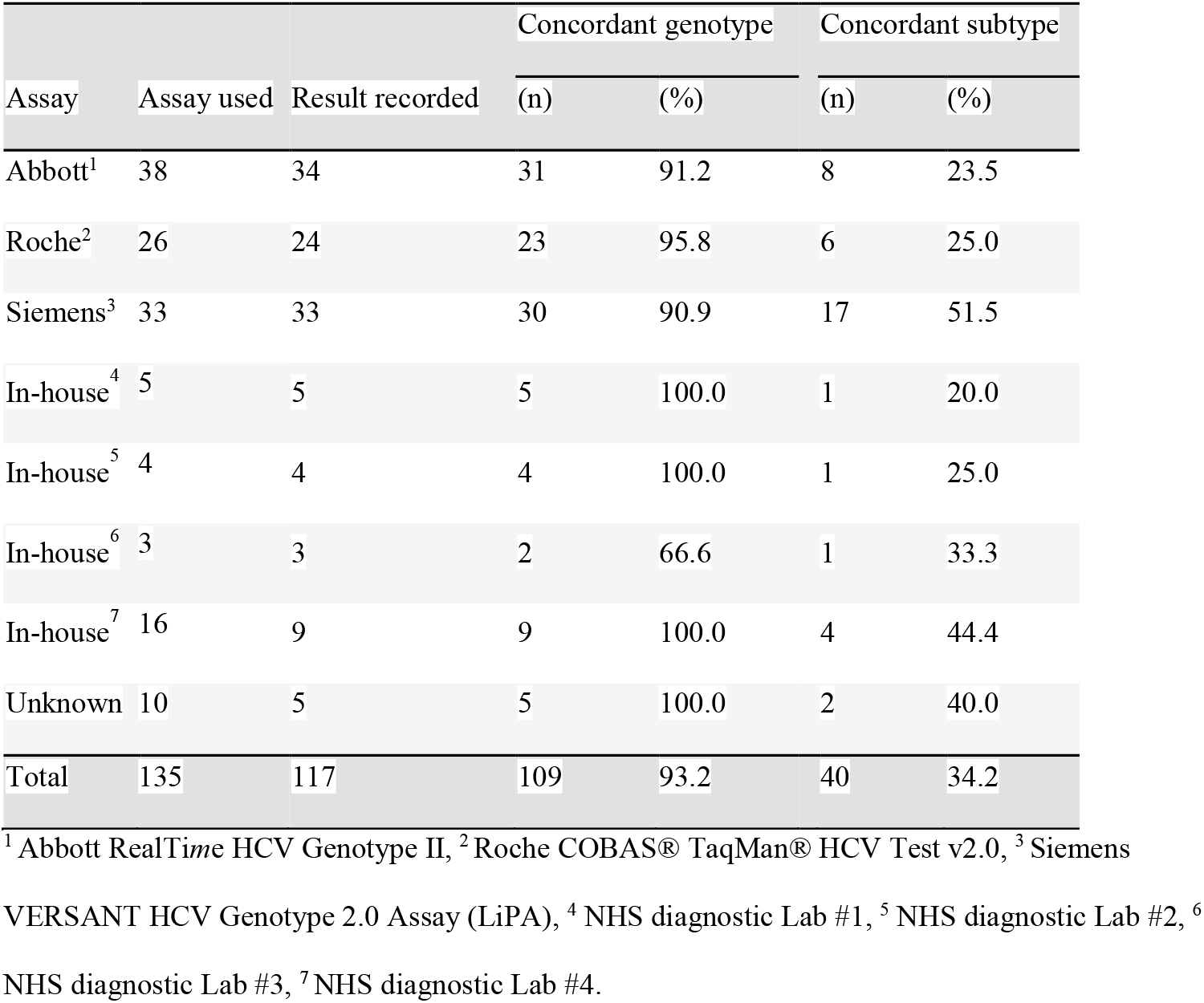
Summary of diagnostic lab subtyping compared to *p*-distance assignment

### Comparison of online HCV genotyping tools

As many diagnostic centres do not have access to commercial genotyping kits regardless of their accuracy, but will be able to request Sanger sequencing of PCR products from commercial laboratories, the partial NS5B sequences were typed using five online HCV tools (Geno2Pheno, Genome Detective, LANL, ViPR, HCV-Glue) and the outputs compared to the subtypes assigned using *p*-distance analysis (**Supplementary Table 5**). 34 samples were discordantly typed compared to the p-distance assigned genotype, or not typed, by at least one of the online tools (**Table 5**). The Geno2Pheno assignments were the most consistent with those determined by *p-*distance with discordant subtypes assigned to 15 samples. However, seven of these can be attributed to the out-of-date reference dataset which does not include the more recently characterised Gt1o. The LANL database, which is no longer curated, produced the most inconsistent subtype assignments (n=26). The majority of the samples (79.4%, n=27) that did not receive a consistent subtype assignment were those with a *p-*distance of ≥0.1 to the closest reference isolate.

**Table 5.** Samples with inconsistent NS5B subtyping by online tools

| Sample ID | *p*-distance subtype | *p*-distance to closest reference‡ | Geno2Pheno | Genome Detective | LANL | ViPR | HCV GLUE |
| --- | --- | --- | --- | --- | --- | --- | --- |
| Cam_01 | 2*/2r | 0.14 | 2j | 2 (unassigned) | 2 (unassigned) | 2 (unassigned) | Unknown |
| Cam_03 | 1o | 0.03 | 1*5 | 1 (unassigned) | 1a/1c | 1c | 1 (unassigned) |
| Con_02 | 4g/4k | 0.13 | 4k | 4 (unassigned) | 4h | 4h | 4h |
| Con_03 | 4r | 0.05 | ✓ | ✓ | 4c/4r | ✓ | ✓ |
| Con_04 | 4b | 0.1 | ✓ | ✓ | 4b/4q | ✓ | ✓ |
| Con_09 | 4k | 0.15 | ✓ | 4 (unassigned) | 4h | 4h | 4h |
| Con_10 | 4k | 0.08 | ✓ | ✓ | 4 (unassigned) | ✓ | ✓ |
| Con_14 | 4r | 0.05 | ✓ | ✓ | 4c/4r | ✓ | ✓ |
| Con_16 | 4c | 0.04 | ✓ | ✓ | 4 (unassigned) | ✓ | ✓ |
| Con_18 | 4c | 0.06 | ✓ | ✓ | 4 (unassigned) | ✓ | ✓ |
| Gam_01 | 1e | 0.07 | ✓ | ✓ | ✓ | 1 (unassigned) | ✓ |
| Gha_01 | 1*/1d | 0.13 | 1*2 | 1 (unassigned) | 1a | Unknown | 1 (unassigned) |
| Gha_02 | 2a | 0.12 | 2*1 | 2 (unassigned) | 2f/2j/2a | 2 (unassigned) | 2 (unassigned) |
| Gha_03 | 1o | 0.16 | 1*2 | 1 (unassigned) | 1c | 1c | Unknown |
| Gha_04 | 1a | 0.14 | ✓ | 1 (unassigned) | ✓ | ✓ | 1 (unassigned) |
| Gha_07 | 2a | 0.12 | 2j | 2 (unassigned) | 2a/2j | ✓ | 2 (unassigned) |
| Gha_08 | 2j | 0.12 | ✓ | 2 (unassigned) | ✓ | 2 (unassigned) | 2 (unassigned) |
| Nig_01 | 1k/1o | 0.14 | 1*5 | 1 (unassigned) | 1c | 1c | 1c |
| Nig_02 | 1d | 0.12 | ✓ | 1h | 1i/1c/1b | 1i/1c | 1h |
| Nig_07 | 4v | 0.04 | ✓ | ✓ | ✓ | 4l/4v | ✓ |
| Nig_09 | 1d/1l | 0.15 | 1c | 1 (unassigned) | 1c | 1 (unassigned) | 1 (unassigned) |
| Nig_10 | 1d | 0.15 | ✓ | 1h | 1a/1b | 1a | 1h |
| Nig_14 | 1o | 0.12 | 1*5 | 1 (unassigned) | 1a/1c | 1 (unassigned) | 1 (unassigned) |
| Nig_16 | 4r | 0.04 | ✓ | ✓ | 4r/4c | ✓ | ✓ |
| Nig_18 | 1o | 0.13 | 1*5 | 1 (unassigned) | 1c | 1 (unassigned) | 1 (unassigned) |
| Nig_20 | 1o | 0.13 | 1*5 | 1 (unassigned) | 1c | 1 (unassigned) | 1c |
| Nig_21 | 1c | 0.16 | ✓ | 1 (unassigned) | 1a/1k | 1c/1a | 1 (unassigned) |
| Nig_23 | 1o | 0.15 | 1*5 | 1 (unassigned) | 1b | 1 (unassigned) | 1 (unassigned) |
| Nig_25 | 1o | 0.12 | 1*5 | 1 (unassigned) | 1c | 1 (unassigned) | 1 (unassigned) |
| Uga_03 | 4*/4v | 0.1 | ✓ | 4 (unassigned) | 4v/4q | 4l/4v/4q | 4 (unassigned) |
| Zam_01 | 1a | 0.03 | ✓ | ✓ | ✓ | unknown | ✓ |
| Zim_03 | 3h | 0.14 | ✓ | ✓ | ✓ | ✓ | Unknown |
| Zim_07 | 3h | 0.15 | ✓ | ✓ | ✓ | ✓ | Unknown |
| Zim_10 | 3h | 0.16 | ✓ | ✓ | ✓ | ✓ | Unknown |
| Different to *p*-distance assignment (total =34) | n/a | n/a | 15 | 21 | 26 | 22 | 24 |

‡, uncorrected *p*-distance

### NS5A subtyping and RAS-typing in SSA samples

To investigate the levels of NS5A RAS present within the cohort, an NS5A gene fragment covering nt 6258-6611 (based on H77 reference clone - AF009606) was amplified and sequenced. Generating NS5A sequence with pan-genotypic primers NS5aFi and NS5aRI was more challenging than for NS5B, with many samples requiring the use of genotype-specific reverse primers to amplify the NS5A region in conjunction with an alternative pan-genotypic forward primer NS5aFO (**Table 1**). It was not possible to amplify NS5A for four samples (Ken_02, Nig_21, Nig_26 and Som_2, subtypes Gt4t, Gt1c, Gt4t and Gt3h respectively, data not shown).

An NS5A *p-*distance matrix was generated (**Supplementary Table 6**) and the subtype determined and compared with the NS5B-derived subtype assignments (**Supplementary Table 7**). The majority of samples (118 of 131, 90.1%,) were assigned the same subtype using NS5B and NS5A sequence data. However, the NS5A region was more variable compared to the NS5B region, with 33 samples having a *p-*distance to closest reference of ≥0.1, with the highest *p-*distance being 0.23 for sample Nig_23. Thirteen samples were assigned different subtypes using NS5B and NS5A (**Supplementary Table 8**). The partial NS5A sequences were analysed using HCV Geno2Pheno to identify any known and potential RAS for six NS5A inhibitors (daclatasvir [DCV], elbasvir [EBR], ledipasvir [LDV], ombitasvir [OBV], pibrentasvir [PIB] and velpatasvir [VEL]) (**Supplementary table 9**). 20 distinct RAS were identified in the samples, including six samples with Y93H. The viruses from these six samples were of subtype Gt1b, Gt3h and Gt4b (two of each). 19 distinct uncharacterised substitutions on scored positions (SoSP) were identified, with 60 samples possessing at least one SoSP (**Supplementary Table 10**). Some SoSP (30Q, 30R, 30S, 30R, 31L, 93N) potentially impact all known NS5a inhibitors.

For OBV and VEL a single uncharacterised SoSP accounted for ∼50% of the total SoSP for that inhibitor (28M and 31M, respectively), whereas there were no predominant SoSP for EBR, LDV and PIB (**Supplementary Table 10**). PIB was associated with the fewest number of uncharacterised SoSPs (n=25) while DCV and OBV were associated with the most (n=64 and 68, respectively). Of greatest concern was that 15 samples had a known or uncharacterised RAS that may impact the efficacy of all classes of inhibitors. It is important to note that for a given DAA, some variants are classified as a RAS irrespective of genotype e.g. Y93H, while some variants are genotype-specific RAS e.g. for DCV, 28M is a RAS only in a Gt4 background [33, 35]. Consequently, of the 29 samples encoding 28M in NS5A, this is only identified as a RAS in 18 samples (all Gt4 infections - **Supplementary table 9**).

All known RAS and uncharacterised SoSP were combined to form a single metric for potential reduced NS5A treatment efficacy and samples grouped by country of patient birth (**Table 6**). This also included sample genotypes for which a drug was not licensed. PIB had the lowest proportion of samples with potentially reduced efficacy (16.8%). At least one drug was predicted to be 100% effective against all of the samples obtained for 11 of 19 countries represented. Worryingly, of the 24 samples from people born in Nigeria, at least 40% had a known or potential RAS affecting one or other of the currently available drugs.

**Table 6.** Samples with known/potential NS5A resistance mutations grouped by country

| Country of birth | Samples‡ (n) | DCV | EBR | LDV | OBV | PIB | VEL |
| --- | --- | --- | --- | --- | --- | --- | --- |
| Angola | 1 | 100.0 | 100.0 | 100.0 | 100.0 | 100.0 | 0.0 |
| Burundi | 2 | 100.0 | 50.0 | 50.0 | 100.0 | 0.0 | 50.0 |
| Cameroon | 4 | 75.0 | 75.0 | 75.0 | 100.0 | 50.0 | 75.0 |
| C.A.R. | 4 | 50.0 | 25.0 | 25.0 | 75.0 | 25.0 | 25.0 |
| Congo | 19 | 57.9 | 31.6 | 31.6 | 47.4 | 0.0 | 5.3 |
| Eritrea | 7 | 71.4 | 14.3 | 0.0 | 71.4 | 0.0 | 0.0 |
| Eswatini† | 1 | 0.0 | 0.0 | 0.0 | 100.0 | 0.0 | 0.0 |
| Ethiopia | 2 | 50.0 | 0.0 | 0.0 | 50.0 | 0.0 | 0.0 |
| Gambia | 1 | 100.0 | 100.0 | 100.0 | 100.0 | 100.0 | 100.0 |
| Ghana | 8 | 87.5 | 87.5 | 87.5 | 87.5 | 25.0 | 37.5 |
| Kenya | 3 | 0.0 | 0.0 | 0.0 | 66.7 | 0.0 | 0.0 |
| Mauritius | 3 | 33.3 | 0.0 | 33.3 | 33.3 | 0.0 | 0.0 |
| Nigeria | 24 | 45.8 | 41.7 | 41.7 | 70.8 | 50.0 | 58.3 |
| South Africa | 6 | 16.7 | 16.7 | 0.0 | 16.7 | 0.0 | 0.0 |
| Somalia | 28 | 57.1 | 17.9 | 17.9 | 67.9 | 7.1 | 10.7 |
| Tanzania | 3 | 0.0 | 0.0 | 0.0 | 33.3 | 0.0 | 0.0 |
| Uganda | 4 | 75.0 | 75.0 | 75.0 | 50.0 | 25.0 | 0.0 |
| Zambia | 1 | 0.0 | 0.0 | 0.0 | 0.0 | 0.0 | 0.0 |
| Zimbabwe | 11 | 54.5 | 45.5 | 45.5 | 72.7 | 9.2 | 27.3 |
| Total | 131 | 53.4% | 53.4% | 32.8% | 64.1% | 16.8% | 22.1% |

†Formerly Swaziland. ‡Does not include four samples not amplified using NS5A assay. DCV, Daclatasvir; EBR, elbasvir; LDV, ledipasvir; OBV, ombitasvir; PIB, pibrentasvir; VEL, velpatasvir.

When the samples were grouped by subtype and analysed for NS5A RAS (**Figure 2**) it was clear that RAS affecting PIB efficacy were almost exclusively found in patients with Gt1. It was also evident that RAS affecting treatment with all NS5A inhibitors were found in the samples of subtype Gt1d, Gt1e, Gt1g, Gt1l and Gt1o. This was also seen to a small extent for subtype Gt1a and Gt3h. The samples of subtype Gt1*/1d and Gt4b (n=3) were predicted to be resistant to all classes of inhibitor except VEL.

**Figure 2.**
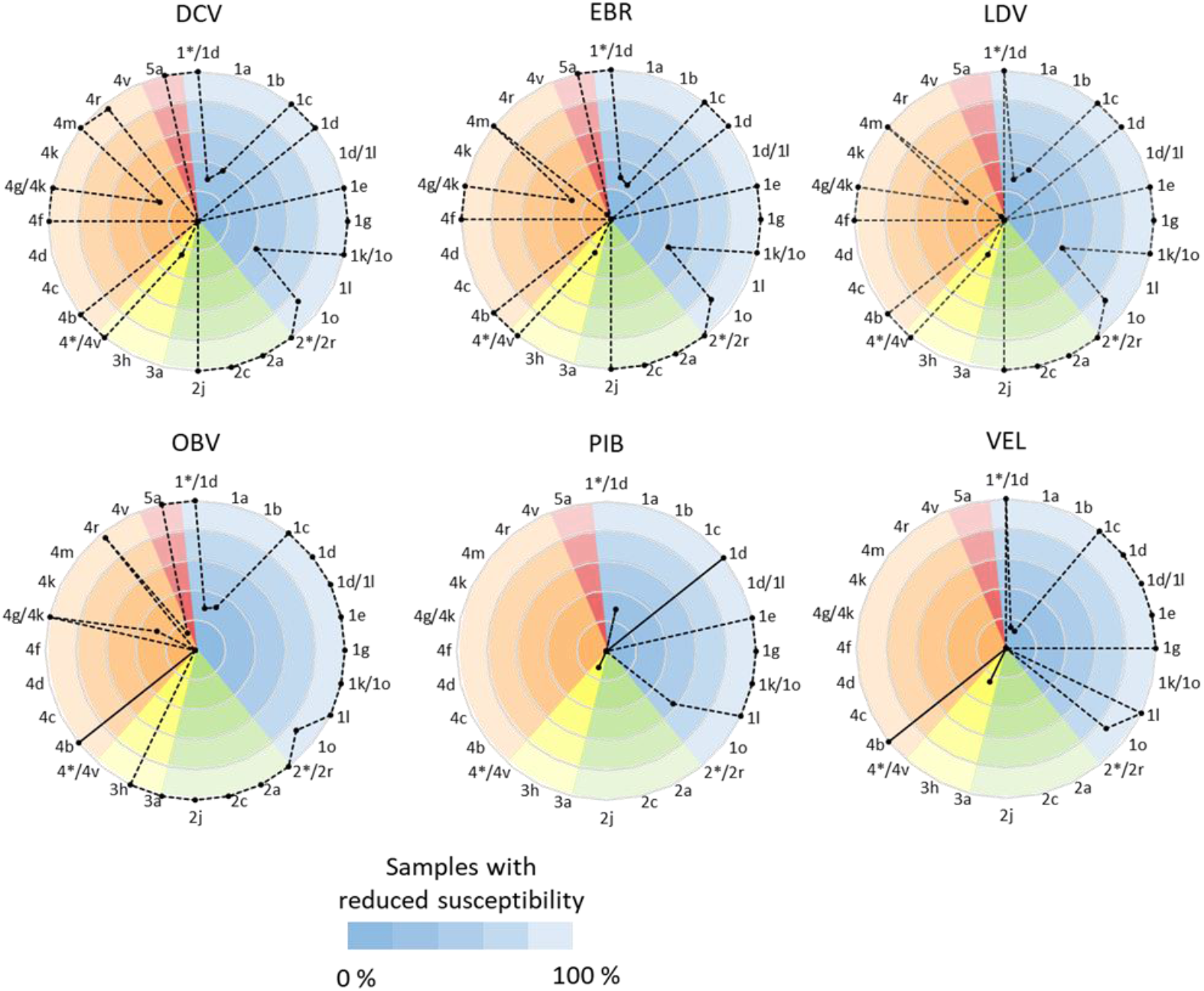
Proportion of SSA HCV subtypes with known or potentially reduced susceptibility to NS5A inhibitors. Radar plots illustrating known of potential reduced efficacy of NS5A inhibitors for treatment of SSA HCV samples. Samples grouped by HCV subtype and clustered by genotype (Gt1, blue; Gt2, green; Gt3, yellow; Gt4, orange; Gt5, red). 6 samples could not be assigned to a single subtype and are shown as duplexed subtypes. Subtypes assessed for their combined predicted response to treatment with a given inhibitor class (0%, all samples fully susceptible; 100%, all samples possessing at least one known or uncharacterised RAS applicable to that drug class). DCV, Daclatasvir; EBR, elbasvir; LDV, ledipasvir; OBV, ombitasvir; PIB, pibrentasvir; VEL, velpatasvir.

## Discussion

Despite the emergence of pan-genotypic DAA regimens, effective treatment and global elimination efforts will require careful surveillance as many first-line therapies are not optimal for all subtypes, especially those circulating in lower- and middle-income countries [4, 36-39]. Even in high-income countries, such as England, not all patients are offered pan-genotypics, especially those with Gt1 infection. In this study we show that commonly used commercial sub-typing assays frequently mistype these non-Western isolates and are essentially of minimal utility when dealing with samples from SSA. In contrast, we show that Sanger sequencing of suitably designed NS5B and NS5A-specific PCR products, allied to basic p-distance or phylogenetic analysis, was able to accurately assign genotype and subtype. Similarly, well-curated and updated online tools, such as Geno2Pheno, Genome Detective and HCV-GLUE all perform considerably better than commercial assays and, in the case of Geno2Pheno and HCV-GLUE, offer additional curated DAA resistance profiles. However, these data were generated using samples containing Gt1-5 HCV, therefore the performance of these assays would benefit from additional validation against Gt6, Gt7 and Gt8 [40-43].

Given the difficulties of obtaining samples in many places with relatively poor health infrastructure, we utilised a large UK HCV study cohort to identify individuals who had most likely acquired HCV whilst living in SSA [22]. This cohort was drawn from the major SSA regions and the resulting analysis highlights the high levels of genetic diversity observed in HCV circulating in this part of the globe and the need for more extensive surveillance.

As part of this enhanced surveillance, there is impetus to deploy next generation sequencing [44]. Whilst this requires significant infrastructural and skills investment that have been addressed to some degree during the COVID-19 pandemic [44], it is still fraught with technical difficulties. For example, many NGS platforms utilise probe- or primer-based enrichment techniques, and the immense genetic diversity of HCV circulating in the field can render these approaches inconsistent. For example, we have recently deployed probe-based capture NGS on samples from the same cohort with variable success [22]. In contrast, we found that a single round PCR assay targeting a partial NS5b gene sequence to be highly effective not only in amplification of extremely diverse isolates, but also in distinguishing their subtypes. When combined with one or more accessible online tools, this will allow any laboratory capable of RT-PCR to correctly subtype their samples in SSA with optional but straightforward ambient shipping of amplicons to a national or international sequencing facility. From a cohort of 146 patients, we successfully amplified NS5B and NS5A PCR products from 135 and 131, respectively. It was unclear why 11 NS5B-reactions failed, although only one sample was known to have a high viral load. Of the NS5B positive samples, the vast majority were successfully amplified using in-house genotype specific NS5a primer sets. Recent studies independently arrived at comparable pan-genotypic NS5A primer sets that target similar conserved genomic regions, but also employing additional measures to improve amplification success, such as inosine degeneracy and utilising higher input serum and RNA [45, 46]. However, the additional nested strategy employed in one method would necessitate an additional level of expertise in molecular contamination control, potentially adding further logistical challenges in most settings [45]. In general, these simple Sanger-based sequencing and subtyping pipelines should be easily deployable in most resource-poor settings [44]. Key features of our described protocol relevant to resource limited settings include ambient-stored lyophilised cDNA reagents, ambient temperature stable *Taq*, purification-free sequencing template preparation, ambient sequence template shipping, and ‘paste and click’ sequence analysis using free online tools. In a parallel study we have also demonstrated that this process can be effectively coupled with dried serum / blood spot samples [47].

In conclusion, there is an urgent need for any nation pursuing HCV elimination to ensure sufficient characterisation and continued surveillance of HCV subtype diversity. Delivery of ineffective DAA treatment risks not only often pressured financial and healthcare resources, but also threatens achieving the agreed public health goals nationally and globally. Continued development of both LMIC healthcare infrastructure and personnel to deploy straightforward and low cost molecular biological techniques such as those described here will enable nations to deliver appropriate anti-HCV therapies.

## Data Availability

All data produced in the present work are contained in the manuscript and supplementary materials and online at GenBank

## Conflict of interest statement

W. L. I. has received speaker and consultancy fees from Roche Products, Janssen-Cilag and Novartis; educational grants from Boehringer Ingelheim, MSD, and Gilead Sciences; and research grant support from GlaxoSmithKline, Pfizer, Gilead Sciences, and Janssen-Cilag.

## Financial support statement

K. A. was supported by the Tertiary Education Trust Fund and the National Institute for Health Research Nottingham Biomedical Research Centre.

## CRediT authorship contribution statement

Kazeem Adeboyejo – Investigation, Data curation, Funding acquisition; Barnabas J. King - Data curation, Methodology, Formal Analysis, Supervision, Validation, Visualization Writing – original draft; Theocharis Tsoleridis - Formal Analysis, Visualization; Alexander W. Tarr - Methodology, Supervision; John McLauchlan – Resources, Project administration; William L. Irving - Resources Project administration, Supervision; Jonathan K. Ball - Conceptualization, Funding acquisition, Supervision Writing – original draft; C. Patrick McClure – Conceptualization, Data curation, Formal Analysis, Investigation, Methodology, Supervision, Validation, Writing – original draft.

All authors - Writing – review & editing

## Supplementary Figure Legends

**Supplementary Figure 1.**
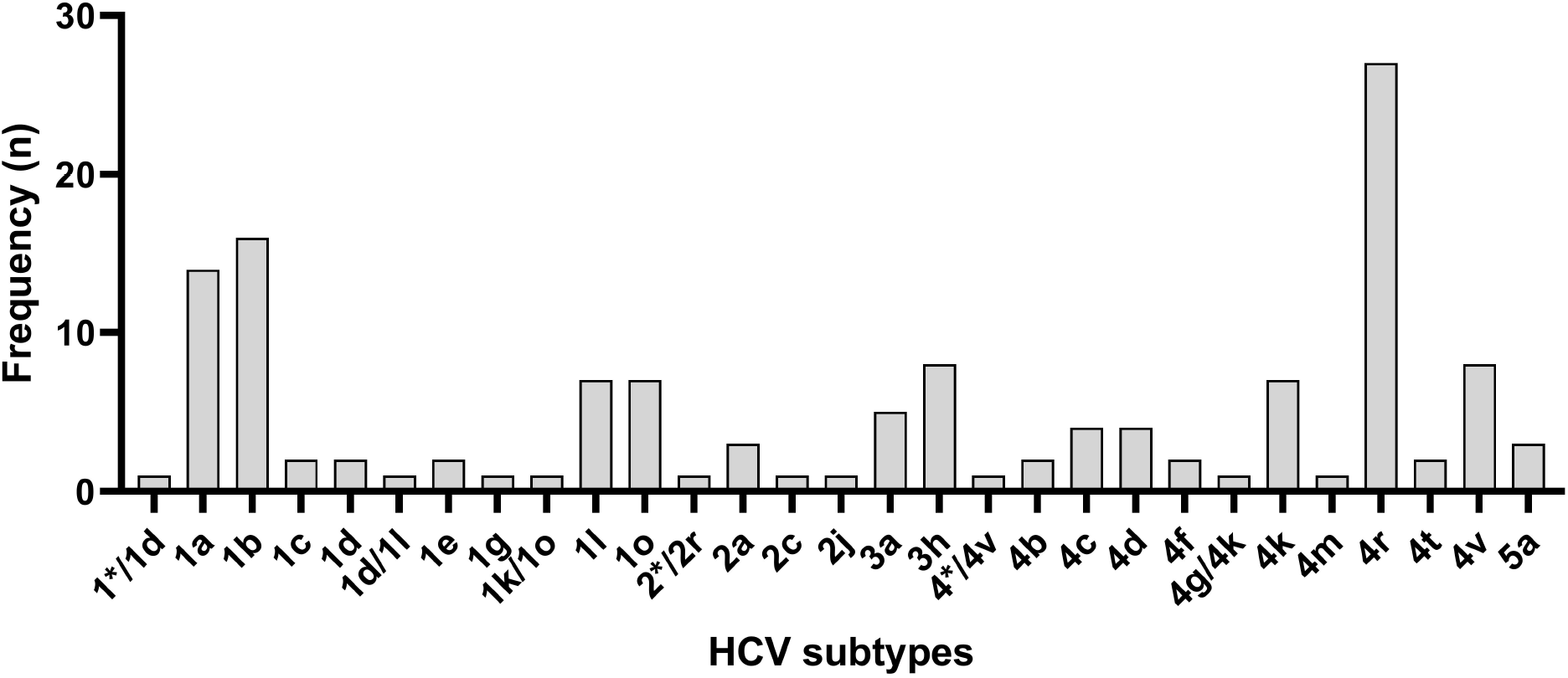
Subtypes identified in samples from patients born in SSA. Samples sub-typed into 23 specific HCV subtypes with 6 samples most closely aligning to isolates from two different subtypes and are shown as duplexed subtypes. Subtypes were determined from *p*-distance matrix of NS5B amplicon and assigned by comparison to closest reference isolate subtype. *undefined subtype.

**Supplementary Figures 2A-F.**
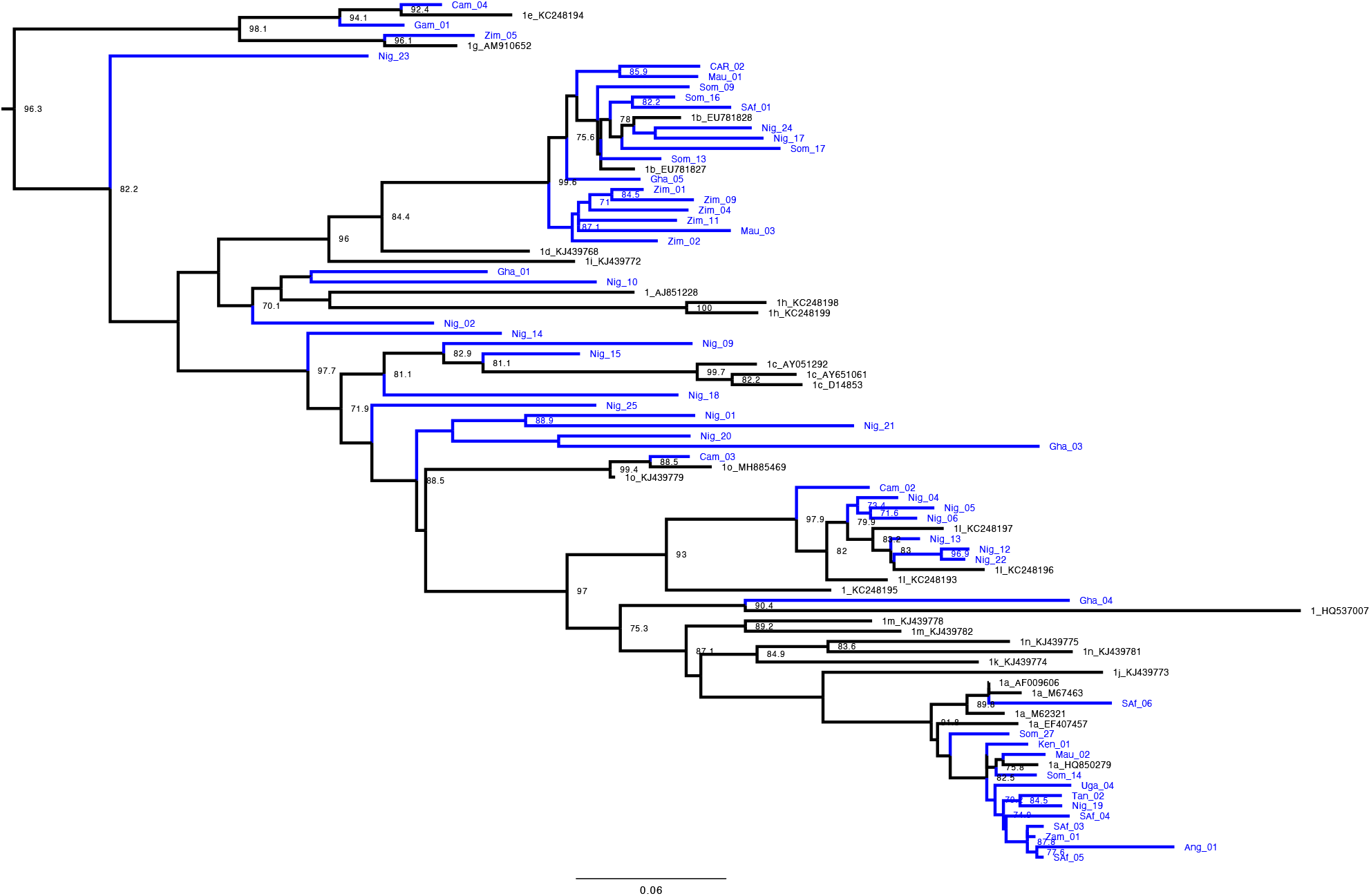
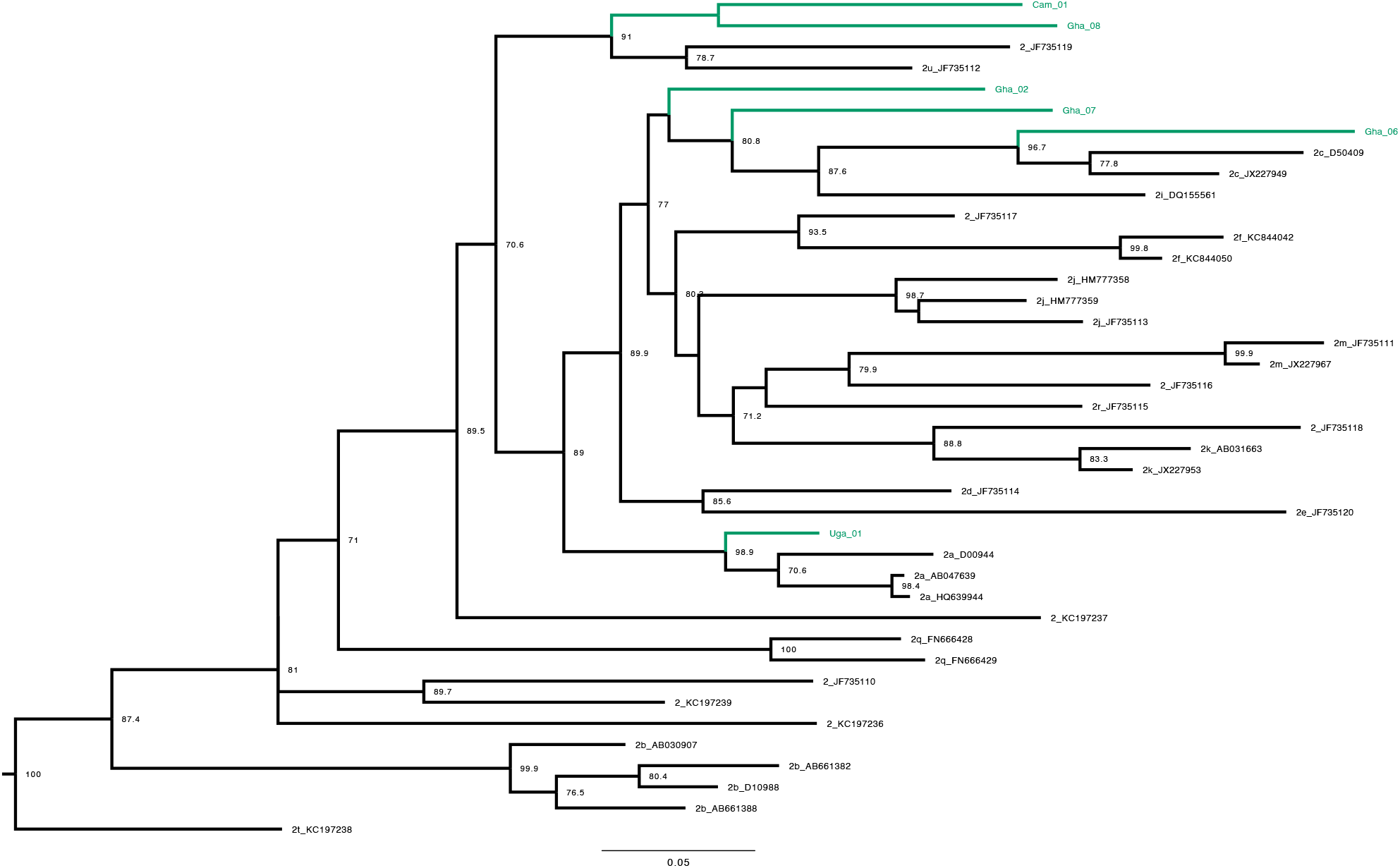
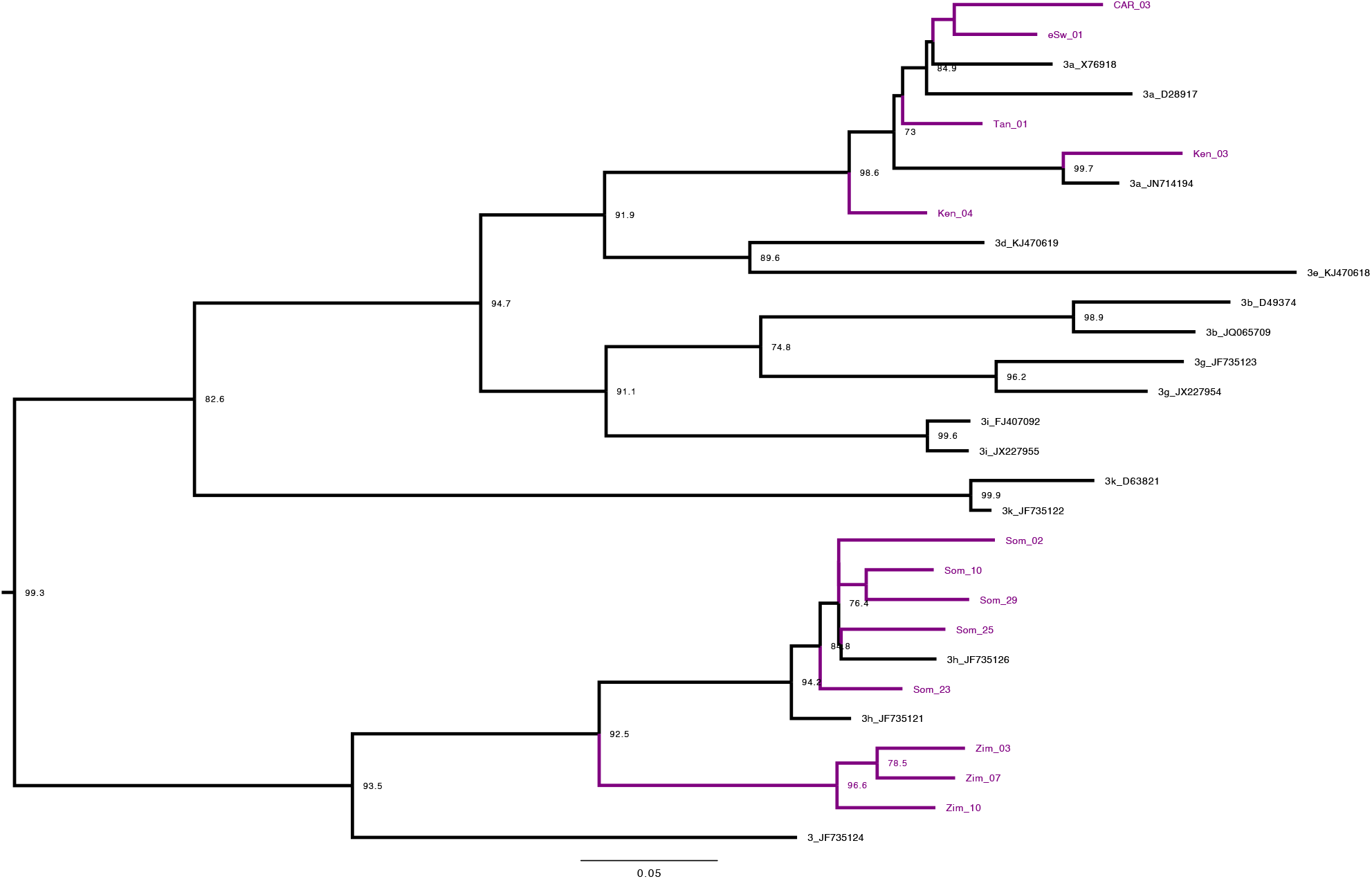
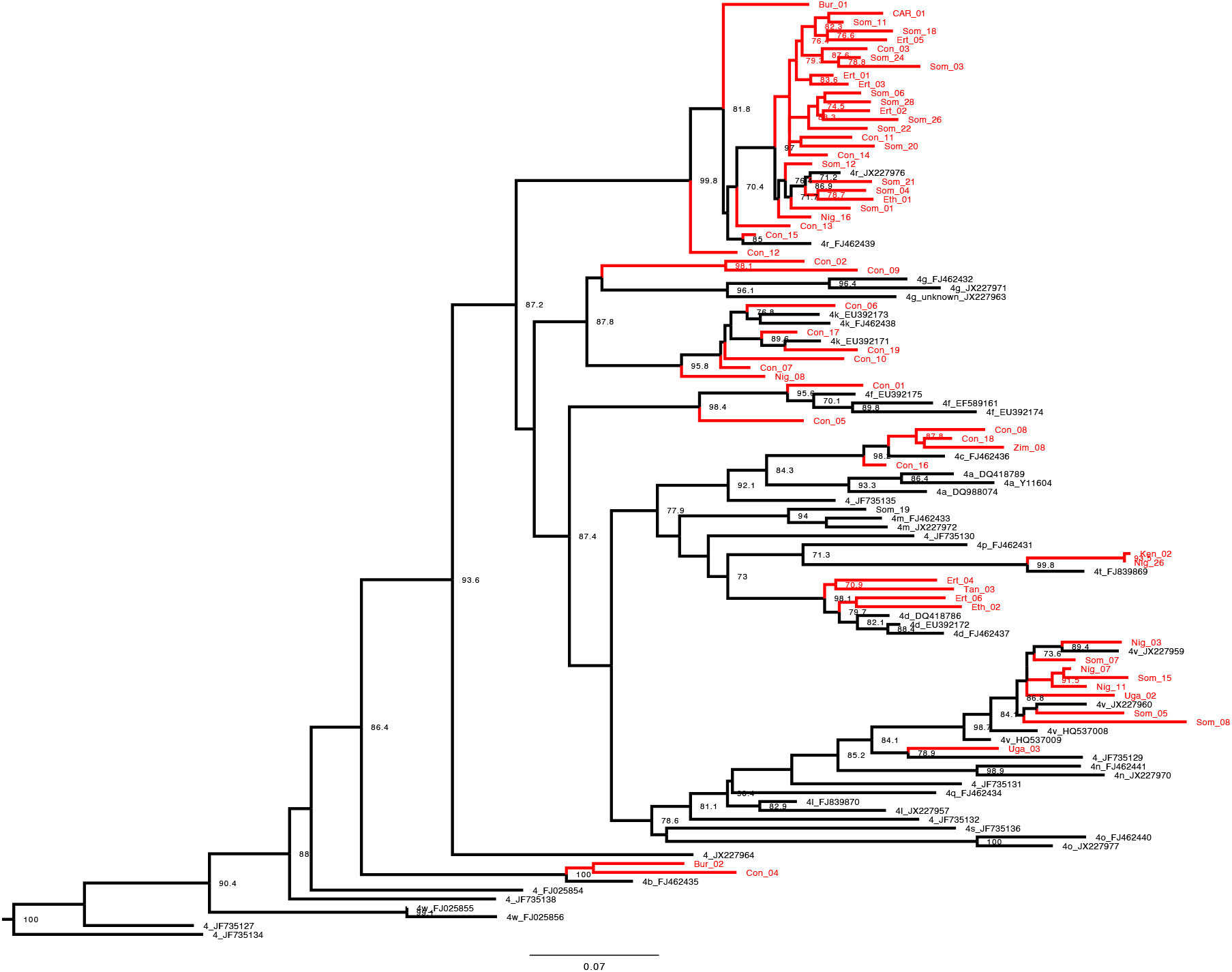
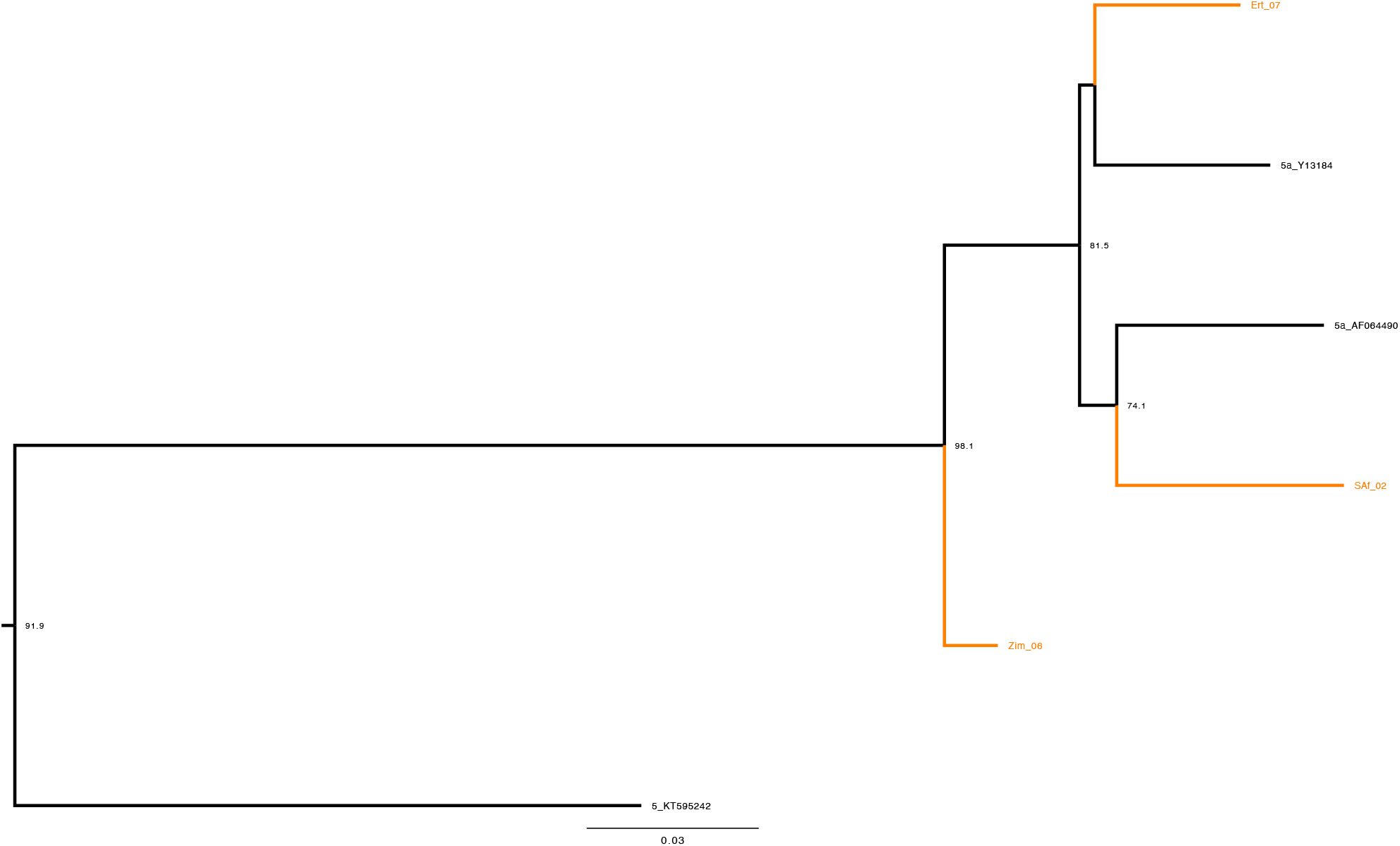
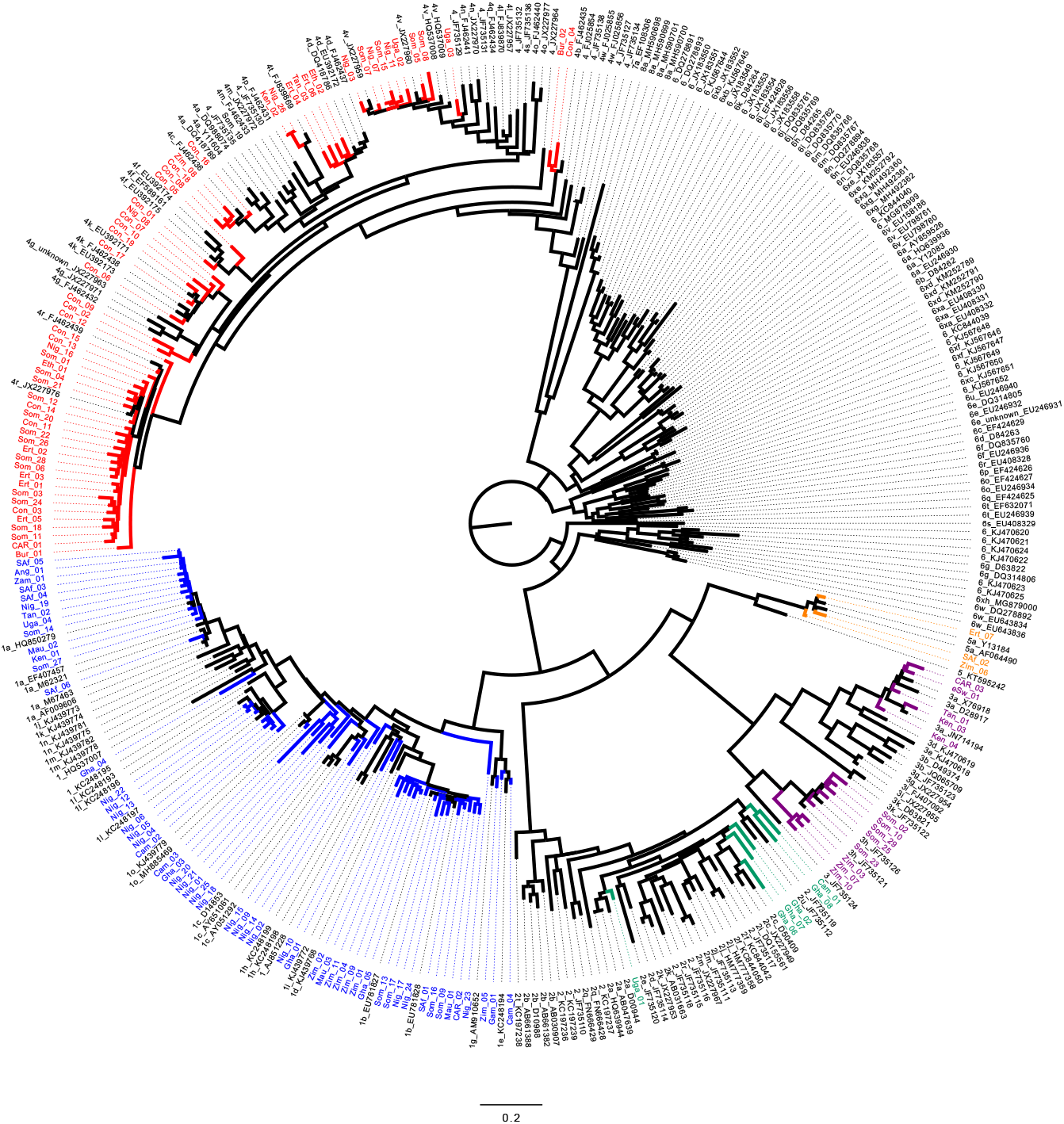
Phylogenetic relationships of HCV sequences based on their partial *NS5b* gene (316nt). The tree represents maximum likelihood phylogenetic analysis of all SSA samples and ICTV reference sequences (352 in total). The SSA samples cluster with their respective genotype reference sequences. Each genotype is coloured uniquely; Gt1 (blue), Gt2 (green), Gt3 (purple), Gt4 (red), Gt5 (orange). Reference sequences are indicated by their GenBank accession numbers. Branch lengths are drawn to a scale of nucleotide substitutions per site. Numbers above individual branches indicate SH-aLRT bootstrap support (only percentages >60 are shown).

## Notes

### Author Declarations

Ethics approval for HCV Research UK was given by National Research Ethics Service (NRES) Committee East Midlands/Derby 1 (Research Ethics Committee reference 11/EM/0314). Use of anonymised samples and associated data for this study was approved by HCV Research UK under reference TR000404.

